# Increased sensorimotor and superior parietal activation correlate with reduced writing dysfluency in writer’s cramp dystonia

**DOI:** 10.1101/2025.03.20.25324331

**Authors:** Noreen Bukhari-Parlakturk, Patrick J. Mulcahey, Michael Fei, Michael W. Lutz, James T. Voyvodic, Simon W. Davis, Andrew M. Michael

**Affiliations:** Department of Neurology, Duke University School of Medicine, Durham, NC 27705, USA; Duke Institute for Brain Sciences, Duke University, Durham, NC 27710, USA; Medical Scientist Training Program, Duke University School of Medicine, Durham, NC 27705, USA; Brain Imaging & Analysis Center, Duke University, Durham, NC 27705, USA

**Keywords:** writer’s cramp dystonia, functional network, sensorimotor cortex, superior parietal cortex

## Abstract

Writer’s cramp (WC) dystonia is a disabling brain disorder characterized by abnormal postures during writing tasks. Although abnormalities were identified in the sensorimotor, parietal, basal ganglia, and cerebellum, the network-level interactions between these brain regions and dystonia symptoms are not well understood. This study investigated the relationship between peak accelerations, an objective measure of writing dysfluency, and functional network (FN) activation in WC and healthy volunteers (HVs). Twenty WC and 22 HV performed a writing task using a kinematic software outside an MRI scanner and repeated it during functional MRI. Group independent component analysis identified 21 FNs, with left sensorimotor, superior parietal, cerebellum, and basal ganglia FNs selected for further analysis. These FNs were activated during writing and no group differences in FN activity were observed. Correlational analysis between FN activity and peak acceleration behavior revealed that reduced activity in left sensorimotor and superior parietal FNs correlated with greater writing dysfluency in WC, a pattern distinct from HVs. These findings suggest that enhanced activation of the left sensorimotor and superior parietal networks may mitigate writing dysfluency in WC. This study provides a mechanistic hypothesis to guide the development of network-based neuromodulation therapies for WC dystonia.

**Author’s summary:** A critical barrier to advancing clinical therapies for writer’s cramp (WC) dystonia is the limited understanding of how brain activation patterns associate with worsening disease severity. Our study addressed this gap by integrating an objective behavioral measure of WC dystonia symptom with changes in functional network activity, revealing the direction of brain activity associated with increased symptom severity. We showed that reduced activity in the left sensorimotor and superior parietal cortices correlated with greater writing dysfluency. These findings suggested that neuromodulation strategies aimed at increasing activity in these cortical regions may offer a promising avenue for developing network-based therapies for WC dystonia.

**Conflict of Interest:** All authors report no financial disclosures or conflicts of interest relevant to this research.

**Authors’ roles:** NBP: conceptualization, data collection, data analysis, statistical analysis, and manuscript writing. PJM: data analysis, and manuscript writing. MF: data analysis. MWL: statistical analysis and manuscript review. JV: study design. SWD: data analysis advice and manuscript critique. AMM: conceptualization, data analysis critique, manuscript writing and critique.

## 1. Introduction

Dystonia is a rare and disabling brain disorder characterized by abnormal postures that can manifest in an isolated limb or be generalized to the whole body (1-3). Despite significant disabilities resulting from this brain disorder, treatment options are limited, with no disease modifying treatments and limited efficacy of symptomatic therapies (1). The absence of disease-modifying treatments for dystonia is partly due to a limited understanding of how alterations in brain activity contribute to the development of dystonia symptoms.

Writer’s Cramp (WC) is a subtype of adult focal dystonia that can be used to study the relationship between brain abnormalities and dystonia symptoms (4). This is because WC dystonia is clinically stereotyped (1, 3), exhibits brain state dependency with controlled writing tasks (4), and symptom severity can be assessed using objective measures of writing dysfluency in such tasks (5).

Prior neuroimaging studies in WC dystonia identified multiple brain abnormalities that spanned the cortical and subcortical brain regions. Specifically, decreased functional activity was reported in brain regions of premotor cortex, parietal cortex, basal ganglia and cerebellum in WC compared to healthy volunteers (HV) (6). Despite knowledge of decreased regional brain activities, uncertainties remain about the relationship between these brain abnormalities and dystonia symptom severity. A barrier to studying the relationship between changes in brain activity and dystonia symptom severity is the lack of objective behavioral measures. Previous studies have used clinician-rated scales and disease duration to measure dystonia symptoms (7, 8). One study demonstrated that increased connectivity between the primary sensory and primary motor cortices was associated with higher scores on a clinician-rated scale in WC while another study reported stronger parietal to premotor connectivity was associated with reduced symptom duration in WC participants (6, 8). Collectively, prior neuroimaging studies in WC identified an association between changes in intra-cortical connectivity and increased symptom severity as assessed by clinician-rated scales or disease duration. It remains unclear whether the changes in brain connectivity to behavior relationship are due to changes in both brain regions or a single brain region.

Furthermore, clinician-rated measures consist of categorical gradations with intra-operator variability that may lack the sensitivity to detect key changes in brain activity and dystonia symptom severity. Recently, a detailed analysis of 22 kinematic writing measures in WC and healthy volunteers (HV) showed that the sum of acceleration peaks in a single sentence (henceforth peak accelerations) demonstrated high diagnostic potential (sensitivity, specificity, and intra-participant reliability) to differentiate WC from HV and associated with patient reported dystonia and disability scales (5). The peak accelerations measure, therefore, represents an objective behavioral measure that can be used to study the relationship between WC dystonia symptom severity and changes in brain activity patterns. Elucidating the relationship between changes in brain activity and dystonia symptom severity may enable us to identify the direction of brain changes that could improve behavior, thereby informing the development of future clinical interventions for this disabling brain disorder.

This study aimed to identify changes in brain activity associated with dysfluent writing behavior in WC. We hypothesized that decreased BOLD activity in sensorimotor cortex may contribute to greater writing dysfluency in WC participants. To test this hypothesis, WC and HV participants completed a kinematic writing assay outside the MRI scanner and then repeated it during a writing task fMRI. Group independent component analysis (ICA) was then used to generate functional networks (FNs), which were subsequently correlated with the behavioral measure of peak accelerations. Overall, this study aimed to investigate the relationship between FN activity and dysfluent writing behavior in WC dystonia to inform future clinical interventions.

## 2. Materials and Methods

### 2.1 Study Design

The study was approved by the Duke Health Institutional Review Board (IRB: 0094131) and performed in accordance with the Declaration of Helsinki. All enrolled participants gave written informed consent. Inclusion criteria for writer’s cramp dystonia were isolated right-hand dystonia during the writing task diagnosed by a Movement Disorder Specialist, more than three months from the last botulinum toxin injection, and more than one month from trihexyphenidyl medication. Exclusion criteria were any contraindications to receiving MRI. Aged-matched, right-hand dominant HVs without structural brain disorders or psychiatric illnesses were also recruited for the study. A total of 44 participants consented to the study (23 HV and 21 WC). All study participants completed a writing behavior assay and a task-based fMRI during two in-person observational research visits.

### 2.2 Writing behavior assay

All participants performed a behavioral writing assay during their first visit. The baseline visit consisted of participants using a sensor-based pen on a digital tablet (MobileStudio Pro13; Wacom Co, Ltd, Kazo, Japan). Participants copied a holo-alphabetic sentence ten times in a writing software (MovAlyzer, Tempe, AZ, USA). The sensor-based pen recorded the x, y, and z positions and the time function of the participants’ writings. The writing software then transformed the writing samples’ position parameters and time functions using a Fast Fourier transform algorithm to calculate the kinematic features automatically, including a measure of peak accelerations. A previously detailed analysis of these kinematic writing measures showed that peak accelerations differentiated WC dystonia from healthy with high sensitivity, specificity, and intra-participant reliability, and associated with patient reported dystonia and disability scales (5).

### 2.3 Brain imaging

All study participants completed a task-based fMRI during their second visit. *<u>Study design:</u>*Participants were visually presented instructions to copy a holo-alphabetic sentence using CIGAL software (9). CIGAL software also recorded the participants’ writing on the tablet (9). Participants copied the sentence in a 20s block design (on-block) alternated by rest blocks of 16s (off-block) for twelve repetitions. To generalize the functional activation pattern during the writing task and minimize learning, participants copied a different holo-alphabetic sentence during each writing block. *<u>Data acquisition:</u>*Structural and functional MRI were acquired from all participants using a 3-Tesla GE scanner. Structural T1-weighted images were acquired using echo-planar imaging with the following parameters: 256 × 256 matrix, TR = 7.316 s; TE = 3.036 s; FOV= 25.6mm^2^, bandwidth 41.67 Hz/Pixel, 1mm slice thickness. Functional echo-planar images were acquired using the following parameters: voxel size = 3.5×3.5×4.0 mm^3^, TR = 2 s, TE = 30 ms, flip angle = 90°, FOV = 22 cm, bandwidth = 250 Hz/Pixel, 37 interleaved slices. During the functional scans, participants were asked to minimize their head movements. *<u>Image preprocessing:</u>* All fMRI images were preprocessed using fMRIPrep, an automated pipeline to perform brain extraction, head motion, distortion, slice timing correction, intraparticipant registration, and spatial normalization (10). fMRIPrep also provided an estimation of scan quality for each fMRI run. Participants with excessive head movements, defined as a mean frame-wise displacement greater than 0.5 mm, were excluded from the study. This threshold resulted in two participants, one from each group, being excluded from further data analysis (final study cohort: 22 HVs and 20 WCs).

### 2.4 Generation of functional networks (FNs)

Pre-processed fMRI were input into GIFT v3.0 Toolbox on MATLAB R2019b to perform group independent component analysis (GICA) (11). Based on the GIFT Toolbox’s estimated number of components across the data set, 46 candidate FNs were generated using the InfoMax algorithm. FNs were then identified by visually comparing them with established brain network atlases (12-15). Of the 46 FNs generated, 21 had spatial distributions consistent with known human brain regions. The FNs for left sensorimotor cortex, superior parietal cortices, basal ganglia-thalamus, and cerebellum were selected for further analysis based on a prior study implicating their role in WC dystonia (6). GICA’s ICASSO function was then used to test the reliability of these known FNs (16). Specifically, GICA was repeated 30 times to compute the Iq index. The Iq index represents high intra-cluster and low extra-cluster reliability. The Iq index was greater than 0.95 for all known FNs, indicating highly reliable, reproducible, and stable FN estimation.

### 2.5 Estimation of FN task activation

Study participant’s FN BOLD timecourses for each of their four FNs were extracted from the GIFT toolbox and z-score normalized across the duration of the writing task. FN activation for the writing task was estimated via the Pearson’s correlation (R-value) between the FN BOLD timecourse and the writing task model timecourse. The writing task model timecourse was computed by convolving the writing task presentation timecourse with FSL’s canonical hemodynamic response function(17). Pearson’s correlations were transformed to Fisher z-scores (18, 19). The approach utilized here is adapted from traditional signal estimation techniques in task fMRI, where the task model time course is compared to the BOLD time course of each voxel (20). In this adaptation, the BOLD time course of each voxel is replaced by the FN BOLD time course.

### 2.6 Statistical analysis

To analyze age and sex differences between HV and WC, an unpaired t-test was performed, and equal variance was not assumed. To evaluate for sex differences, a Fisher exact test was performed. To analyze for group differences in MRI head motion parameters and peak accelerations behavior, an unpaired t-test was performed, and equal variance was not assumed. To analyze group differences in BOLD activity in each FN, a mixed effects model for repeated measures was performed. Changes in BOLD activity during the on-block and off-block of writing were analyzed separately. The dependent variable was BOLD activity for each FN. The covariates were group (HV vs. WC), and repeated measures were TR with compound symmetry covariance structure. To analyze for group differences in FN correlation to the writing task time course, the Fisher z-scores between the two groups in each FN were compared using an unpaired t-test and equal variance was not assumed. To analyze the correlation between FN BOLD activity and peak accelerations behavior, each participant’s Fisher z-scored FN BOLD activity during the writing task was correlated with their peak accelerations’ behavior and presented on a scatterplot graph with the R-value for each group. To analyze for group differences in correlation of FN BOLD activity to peak accelerations behavior, a generalized linear regression analysis was performed. The dependent variable was peak accelerations behavior, the covariates were group (HV vs. WC), FN activity and interaction term (group*FN activity). All generalized linear regression results were adjusted for multiple comparisons using the Benjamini-Hochberg method (17, 21).

## 3. Results

### 3.1 Group characterization

Data from forty-two participants (22 HV and 20 WC) were used in the present study. There were no significant differences in mean age (t(39)= −0.354, p = 0.725) or sex (p = 0.315, Fisher exact test) (**Table 1**). All WC participants reported onset of dystonia symptoms during the task of writing. Six of the 20 participants in the study reported dystonia symptoms only occurred during the task of writing (simple writer’s cramp) while 14 of the 20 participants reported dystonia symptoms occurred with other tasks in addition to writing (complex writer’s cramp). The mean duration of dystonia symptoms was 16.2 years [SD 14.10]. A comparison of kinematic writing behavior demonstrated that individuals with WC manifested greater peak accelerations behavior during writing compared to HV (HV mean: 308 [SD 52] counts, WC mean 472 [185] counts, HV vs. WC mean difference: 164 counts, t(22): 3.83, p = 0.0009, unpaired t-test) **(Figure 1)**.

**Table 1:** Study participant demographics.

|  | HV<br>(n = 22) | WC<br>(n = 20) | p-value |
| --- | --- | --- | --- |
| Age (years) | 57 [12.93] | 55 [13.79] | 0.725 |
| Sex (M/F) | 14/8 | 16/4 | 0.315 |
| Disease duration (yrs) |  | 16.2 [14.10] |  |

Data are presented as mean [SD].

**Figure 1.**
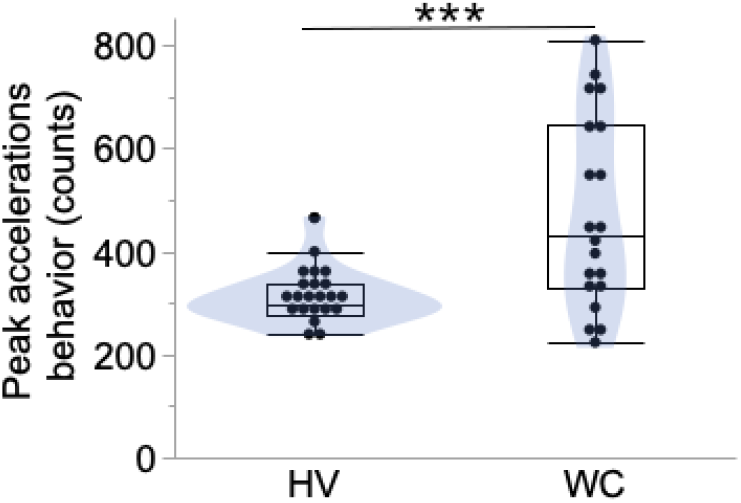
WC participants show greater dysfluent writing behavior. Participants copied ten sentences in a digital software that reported the number of peak accelerations in each sentence. Each data point represents the mean peak acceleration counts per participant with box plots showing the 25%, 50% and 75% of the data distribution and the error bars showing the minimum and maximum. ***p<0.001

### 3.2 No differences in MRI head motion parameters

To investigate the brain changes during the dystonia inducing writing task, participants performed a sentence copying task in a block design (**Figure 2A**). We compared the head motion parameter of mean frame wise displacement between HV and WC participants during the writing task. There were no significant differences in mean frame wise displacement across the two groups (HV: 0.20 [0.07] a.u., WC: mean 0.26 [SD 0.12] a.u., p = 0.10, two tailed unpaired t-test). Therefore, BOLD activity of the four FNs across the two study cohorts was technically comparable.

### 3.3 Comparable FN engagement during dystonic writing task

Preprocessed fMRI brain images during the writing task were input into a GICA analysis to generate FNs (**Figure 2A**). Of the 21 FNs with identifiable brain regions generated by GICA, the FNs for left sensorimotor cortex (SM-L), superior parietal cortices (SPC), cerebellum (CBL) and basal ganglia (BG) were selected for further analysis (**Figure 2B-E**). As expected, the time courses of these FNs demonstrated increased activation during the writing task and decreased activation during the rest period. A comparison of the activation patterns between WC and HV revealed no statistically significant differences in activation during the writing or rest blocks (**Table 2**). However, trends were observed during the writing block with WC demonstrating decreased SML activation (difference − 0.08 a.u., p = 0.073) and decreased basal ganglia activation (difference = −0.16 a.u., p =0.069) compared to HV. During the rest block, WC demonstrated a trend towards increased basal ganglia activation in WC compared to HV (difference = 0.18 a.u., p = 0.09).

To determine if there were any delays in FN activation relative to the writing task presentation, each FN in HV and WC was correlated with the writing task model timecourse and there were no group differences observed (SML: HV vs. WC mean difference = − 0.02 a.u., p = 0.787; SPC: − 0.15 a.u., p = 0.168; CBL: − 0.07 a.u., p = 0.404; BG: −0.13 a.u., p = 0.223).

**Figure 2.**
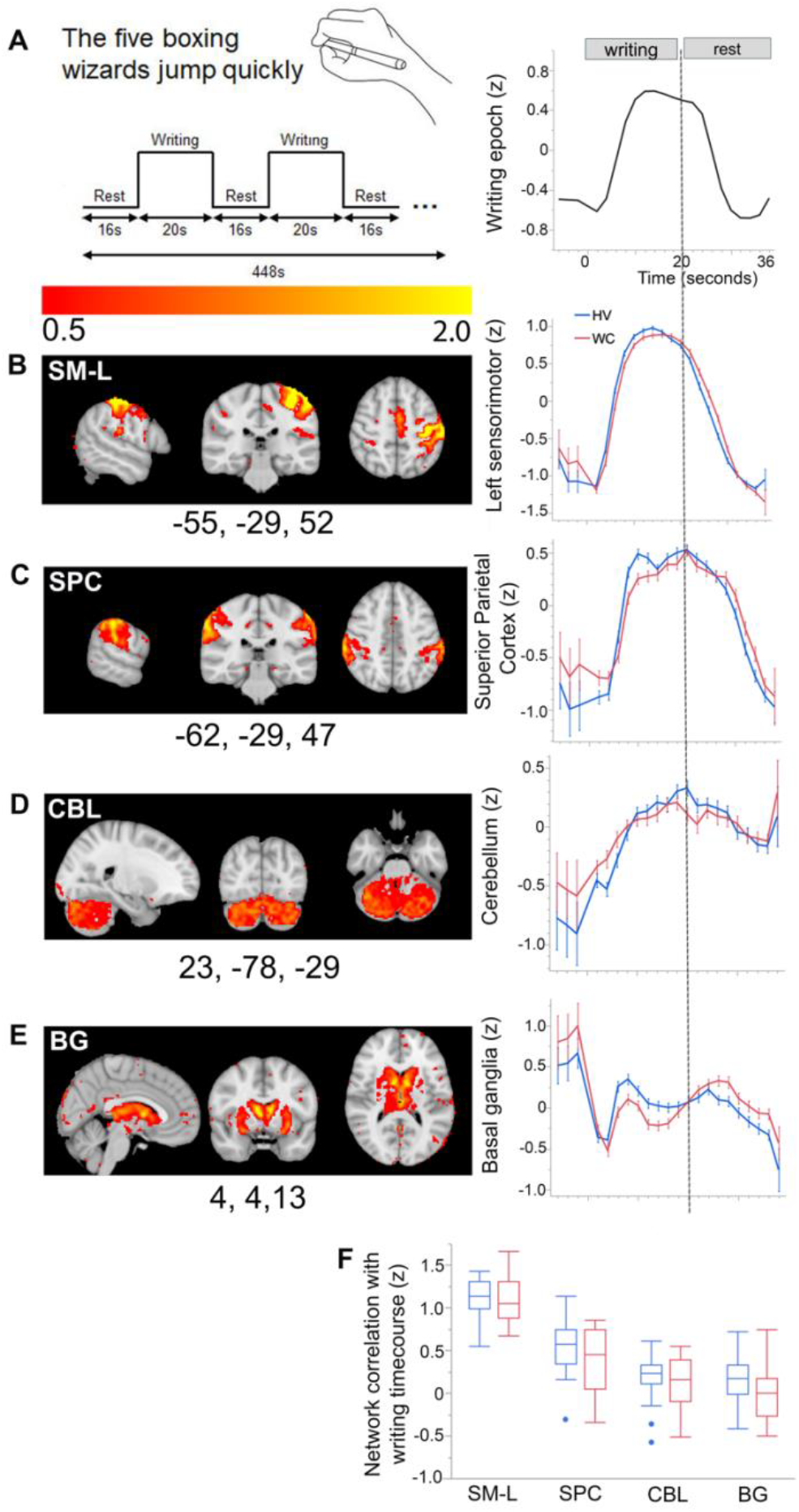
WC and HV participants’ functional networks (FNs) show comparable engagement to the writing task. **A)** Participants performed sentence copying task alternated by rest in MRI scanner for twelve writing epochs. GICA analysis was used to generate FNs **(B-E, left panel)**. Group differences in FN BOLD activity during the writing epoch were compared for **B)** left sensorimotor (SM-L), **C)** bilateral superior parietal cortices (SPC), **D)** cerebellum (CBL) and **E)** basal ganglia (BG). **F)** The correlation between the writing task model timecourse and FN BOLD activity in HV (blue lines and boxplot) and WC (red) were evaluated. There were no statistical differences between HV and WC in activation of the four networks during the writing task.

### 3.4 Decreased SML and SPC FN activity correlates with increased dysfluent writing

To investigate the relationship between changes in FN activity and dysfluent writing behavior, each FN’s mean BOLD activity was correlated with peak accelerations across all participants. WC participants showed a strong correlation between increasing SML activity and decreasing peak acceleration behavior. This correlation was significantly different from HV (WC: R= −0.67, p =0.001; HV: R= −0.28, p = 0.22, HV vs. WC mean difference: 656, p = 0.006, generalized linear regression) (**Figure 3A and Table 3)**. WC participants also showed a strong correlation between increasing SPC activity and decreasing peak acceleration that was significantly different from HV (WC: R= −0.65, p=0.002; HV: R = −0.10, p = 0.66 HV vs. WC mean difference: 467, p = 0.006, generalized linear regression)) (**Figure 3B**). No significant correlations were observed between FNs of BG and CBL and peak accelerations behavior in HV or WC (**Figure 3C and 3D** and **Table 3**).

Overall, a brain-behavior correlational analysis revealed that SML and SPC FN activation inversely correlated with dysfluent behavior in WC but not HV suggesting that increasing activation of these two FNs selectively correlates with reduced dysfluent behavior in WC participants.

**Table 2:** Differences in functional network (FN) activation during writing and rest blocks in healthy and writer’s cramp dystonia.

| FNs | Task block | HV | WC | Mixed model for repeated measures |  |
| --- | --- | --- | --- | --- | --- |
|  |  | Mean (SE) |  | HV vs. WC difference (SE) | p-value |
| SML | writing | 0.45 (0.032) | 0.37 (0.033) | -0.08 (0.046) | 0.073<sup>t</sup> |
| SML | rest | -0.44 (0.054) | -0.33 (0.057) | 0.11 (0.078) | 0.157 |
| SPC | writing | 0.18 (0.045) | 0.10 (0.047) | -0.08 (0.065) | 0.240 |
| SPC | rest | -0.02 (0.056) | 0.07 (0.059) | 0.09 (0.082) | 0.288 |
| CBL | writing | 0.07 (0.055) | 0.05 (0.058) | 0.02 (0.080) | 0.814 |
| CBL | rest | 0.06 (0.068) | 0.04 (0.071) | -0.03 (0.098) | 0.794 |
| BG | writing | 0.07 (0.058) | -0.09 (0.061) | -0.16 (0.084) | 0.069<sup>t</sup> |
| BG | rest | 0.003 (0.070) | 0.18 (0.074) | 0.18 (0.102) | 0.090<sup>t</sup> |

SE: standard error; p-values are unadjusted. t = trend towards $p < 0.05$

**Table 3:** Association between functional network (FN) activation during writing task and peak accelerations behavior in healthy and writer’s cramp dystonia.

| Functional Networks | HV |  | WC |  | Generalized linear regression |  |
| --- | --- | --- | --- | --- | --- | --- |
|  | R | p-value | R | p-value | HV vs. WC mean difference (SE) | p-value |
| SML | -0.28 | 0.58 | -0.67 | 0.008 | 656 (193) | 0.006 |
| SPC | -0.10 | 0.81 | -0.65 | 0.008 | 467 (147) | 0.006 |
| CBL | -0.08 | 0.81 | -0.10 | 0.81 | 42 (159) | 0.48 |
| BG | -0.06 | 0.81 | -0.22 | 0.72 | 137 (148) | 0.79 |

SE: standard error; p-values adjusted for multiple comparison at $p < 0.05$

| Functional Networks | HV |  | WC |  | Generalized linear regression |  |
| --- | --- | --- | --- | --- | --- | --- |
|  | R | p-value | R | p-value | HV vs. WC<br>mean difference (SE) | p-value |
| SML | -0.28 | 0.58 | -0.67 | 0.008 | 656 (193) | 0.006 |
| SPC | -0.10 | 0.81 | -0.65 | 0.008 | 467 (147) | 0.006 |
| CBL | -0.08 | 0.81 | -0.10 | 0.81 | 42 (159) | 0.48 |
| BG | -0.06 | 0.81 | -0.22 | 0.72 | 137 (148) | 0.79 |

SE: standard error; p-values adjusted for multiple comparison at $p < 0.05$

**Figure 3:**
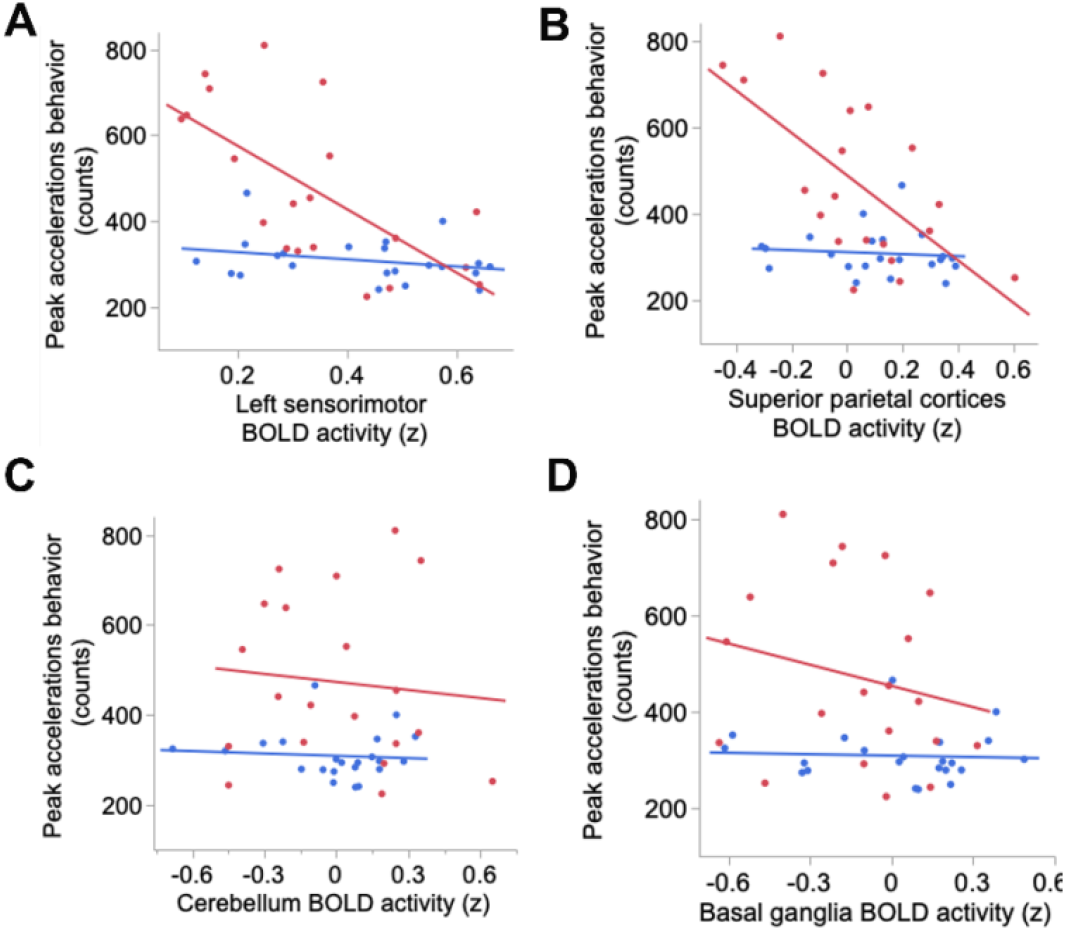
Increased activity in left sensorimotor and superior parietal functional networks (FNs) correlated with reduced writing dysfluency in WC dystonia. Each data point represents a participant’s correlation between their FN activity and peak accelerations behavior. Increased activation of the left sensorimotor and superior parietal FNs correlated with decreased peak acceleration behavior in WC (red line), distinct from HV (blue).

## 4. Discussion

The study aimed to understand the relationship between FN activity and dysfluent writing behavior in WC dystonia. The study employed a three-pronged methodological approach. First, we employed a data driven analysis to organize brain regions into temporally coherent FNs and selected four FNs based on their relevance to to the pathophysiology of dystonia. Second, we used a software-based objective writing metric with high sensitivity and reliability, overcoming the limitations of clinician-rated scales. Thirdly, the study incorporated a within-participant design wherein participants underwent behavioral and fMRI assessments, allowing for an examination of the neural correlates of dystonic symptoms at the individual level. The key study finding is that reduced BOLD activity in the left sensorimotor and superior parietal FNs correlated with increased dysfluent writing behavior. Therefore, clinical interventions that increase activation of these brain regions may reduce dysfluent writing behavior in WC. Findings from this study provide a mechanistic hypothesis to guide the development of neuromodulation therapies for focal hand dystonia.

The primary study finding is that decreased activation of the left sensorimotor FNs is associated with increased writing dysfluency in WC. Writing is a learned motor skill that requires receiving sensory input from the hand and precisely executing the hand movements. In healthy participants, right handwriting activates the left premotor cortex, a subregion of the sensorimotor cortex (22). In contrast, right handwriting in WC demonstrated decreased BOLD activity in the left premotor cortex (6). The present study affirmed this finding by demonstrating a trend towards decreased BOLD activity in the left sensorimotor FN. Notably, the present study builds upon prior research by demonstrating that decreased BOLD activity in the left sensorimotor FN correlated with greater disease severity in WC dystonia.

Another key study finding was that decreased BOLD activity in the superior parietal cortex correlated with increased writing dysfluency in WC. The superior parietal cortex plays a crucial role in generating a mental representation of limb function and integrating complex sensory information to guide precise limb movements, a process known as sensorimotor integration (23, 24). Reduced BOLD activity and connectivity in the superior parietal cortex was previously reported in WC (6, 25). Increasing parietal-premotor cortex connectivity correlated with reduced disease severity as measured by a dystonia rating scale (6, 25). However, it remained unclear if this brain connectivity to behavior relationship was due to changes in both parietal and premotor cortex activity or only one of these brain regions. The present study suggests that a functional impairment in superior parietal cortex activity contributes to greater dystonia disease severity in WC.

Our study findings have important implications for clinical therapies for WC dystonia. The inverse correlation between the left sensorimotor and superior parietal cortex activity and dysfluent writing behavior suggests that increasing BOLD activity in these brain regions may alleviate dystonia symptoms. Non-invasive brain stimulation techniques, such as transcranial magnetic stimulation (TMS) could be used to increase activation of these cortical brain regions. To date, two studies applied TMS to the primary somatosensory cortex and reported behavioral benefit in WC participants (26, 27). One prior study applied TMS to the inferior parietal lobe and reported increased parietal-premotor cortex connectivity in WC (28). The present study findings align with prior TMS studies reporting increased activation of primary somatosensory and parietal cortices may improve behavioral outcome or brain connectivity in WC dystonia.

The present study also observed trends towards decreased basal ganglia activity during the writing task. These findings align with prior studies reporting decreased BOLD activity in the left putamen, a subregion of the basal ganglia, in WC participants (6). Interestingly, our study also found a trend toward increased BG activation during rest in WC. Future studies with larger samples sizes are needed to investigate this reciprocal brain activation pattern in the basal ganglia during the active writing and rest brain states in WC participants.

While this study provides valuable insights, several limitations should be acknowledged. First, although the sample size was sufficient to detect significant brain-behavior relationships, it may have lacked the statistical power to detect smaller, yet potentially meaningful, group differences in FN activation. Future studies with larger cohorts are, therefore, needed to confirm these study findings and explore additional FN activity to behavior relationships. Second, the study focused on a single writing task, which may not capture the full spectrum of dystonic symptoms in WC. Including other tasks that elicit dystonia, such as typing or music playing, could provide a more comprehensive understanding of the neural mechanisms underlying task-specific focal hand dystonia. Lastly, a theoretical consideration in neuroimaging studies is that network-level changes observed during motor tasks may be confounded by differences in movement performance rather than underlying brain pathology. However, our findings argue against this possibility, as we did not observe the same correlation between FN activity and behavior in both healthy controls and dystonia participants.

In summary, a critical knowledge gap in the field of dystonia neuroimaging is the limited understanding of the complex relationships between brain abnormalities and behavioral manifestations of dystonia, which ultimately hampers the development of more effective clinical therapies for individuals with dystonia. Our study attempts to bridge this gap by coupling objective writing measures with a FN-based brain imaging analysis to demonstrate that reduced activity in the left sensorimotor and superior parietal cortices contributes to greater dysfluent writing behavior in WC participants. These findings also suggest that enhancing activity at the left sensorimotor and superior parietal cortices may improve dystonic writing symptoms in WC and provide a mechanistic hypothesis for the development of targeted neuromodulation therapies for this debilitating disorder.

## Data Availability

All data produced in the present study are available upon reasonable request to the authors

## Acknowledgment

This work was supported by grants to NBP from Dystonia Medical Research Foundation (Clinical Fellowship Training Program), Doris Duke Charitable Foundation (Fund to Retain Clinician Scientists), American Academy of Neurology (career development award) and NIH NCATS (1KL2TR002554). NBP was also supported by a career development award from the Dystonia Coalition (NS065701, TR001456, NS116025) which is part of the National Institutes of Health (NIH) Rare Disease Clinical Research Network (RDCRN), supported by the Office of Rare Diseases Research (ORDR) at the National Center for Advancing Translational Science (NCATS), and the National Institute of Neurological Diseases and Stroke (NINDS). The content is solely the responsibility of the authors and does not necessarily represent the official views of the funding agencies.

The authors would like to thank Tiffany Tran, Amber Holden, Ashley Pifer, and Kelsey Ling, who served as Clinical Research Coordinators for this study, and Mariusz Derezinski-Choo, who assisted with fMRI preprocessing. The authors would also like to thank Dr. Nicole Calakos and Dr. Scott Huettel for their input on data analysis and constructive feedback on the manuscript drafts.

